# An Evidence-Based Methodological Framework for Pandemic Preparedness to Support the Clinical Trial Unit Workforce

**DOI:** 10.1101/2023.03.17.23287311

**Authors:** Peter Phiri, Jian Qing Shi, Heitor Cavalini, Lucy Yardley, Katharine Barnard-Kelly, Sana Sajid, Vanessa Raymont, Shanaya Rathod, Gayathri Delanerolle

## Abstract

**Aims:** This study reports a new workforce preparedness framework for use during pandemics, specifically within clinical trials units.

**Methods:** An evidence-based framework was developed using qualitative and quantitative data, as reported by the EPIC observational study. A framework methodology was used to analyse qualitative and quantitative data to identify themes. The themes were used to identify sub-themes that were codes with illustrative quotes. A logic model was develop using spatial features.

**Results:** The qualitative component of the study included the views of 6 semi-structured interviews where discussions indicated the need for flexible working, requirement for better operational management, and access to electronic data systems remotely.

**Conclusion:** Significant mental health impact on the CTU workforce can be prevented by the introduction of a framework to streamline operational delivery of research, providing flexible working patterns to the workforce, and improved access to health and wellbeing practices. Funding calls should be made available to conduct further workforce-based research in the UK and to develop evidence-based policies to better prepare for future pandemics.

What is already known?

- Epidemic preparedness data indicate many countries remain unprepared
- There are large gaps in knowledge and practice base for continuity of research conduct during a lock-down circumstances
- Generic pandemic preparedness frameworks were available although these had limited relevance to Higher Education Institutions (HEIs) and National Health Services (NHS) that conduct significant volumes of clinical research studies

What are the new findings?

- Epidemic Preparedness Index (EPI) that uses a ranking approach in 188 countries have been developed. The EPI includes health capacities and capabilities, including non-healthcare system features
- The use of EPI scores to correlate with proxy measure for preparedness including detection, investigation and reporting of any outbreaks as well as vaccination rates. Examples include the UK flue vaccination rates and the 2009 H1N1 influenza pandemic
- Capacity to detect and respond to epidemics and pandemic appear to be weak across a number of global regions including Asia and Africa despite the higher risk of emergence of pathogens
- Impact of a pandemic on the healthcare and clinical research workforce is significant. There are many limitations in terms of the support available to manage their own wellbeing
- Different levels of complexities exist globally in terms of research regulations and legislations which impact the efficiency of setting up and conducting a study
- The impact of the pandemic to clinical trial unit staff in the UK indicated a number of aspects that need to be improved pertinent at an organisational and individual level

What do the new findings imply?

- Healthcare and Academic institutions as well as the internal units require fit-for-purpose preparedness procedures
- Improved risk planning and mitigation frameworks would be required to better understand and develop methods to continue to deliver work

## Introduction

Pandemics are a natural communicable disease phenomenon caused by the emergence and spread of a pathogen. The SARS-CoV-2 is the pathogen that led to the COVID-19 pandemic which accelerated global morbidity and mortality. COVID- 19 crippled the world with its ability to transform and transmit relatively rapidly given the patterns of incidence have been different in comparison to other communicable diseases. Wealthier countries endured a significant burden in comparison to low-income countries. High- and middle- income countries constitute to 48% of the global population yet contributed to 53% of the estimated excess mortality-adjusted cumulative deaths due to COVID-19 in 2021 despite having a higher vaccination rate since December 20202, in comparison to those in low and lower-middle income countries [1,2]. Epidemiologically, this is an interesting facet to consider in terms of workforce impact for those working in clinical research.

Rapid transmission rates resulted in global healthcare systems and their workforce to be put under considerable pressure. A study conducted in China at the early stages of the COVID-19 pandemic showed healthcare workers had a high prevalence of anxiety and post-traumatic stress disorder, especially among female front-line staff in Wuhan [3]. These results were replicated in similar study conducted in the UK [4,5]. Further research has shown healthcare workers showed a significant mental health impact in comparison to patients [6]. The mental health impact among healthcare workers were significantly poorer during COVID-19 in comparison to the previous MERS and SARS pandemics [7]. Healthcare workers’ work-related stress elevated the risk of psychological outcomes. In the UK, healthcare workers have highly demanding workloads with the pandemic exacerbating this to further, increasing risks of burnout, moral injury and poor mental health outcomes. The consensus on the wellbeing and mental health of the global population 2 years on from the pandemic has been adversely affected but at varying degrees among different populations. Important predictors of poor mental health have been the disruption and loss of income as well as prolonged illness. Much of the workforce research to assess the mental health impact was focused on clinical staffing groups. Comprehensive longitudinal data is lacking and would be required to show long-term effects of the pandemic among all healthcare groups dispersed within hospital and academic organisations. As governments continue to act as the first line of response to pandemics, resources within healthcare and academic institutions’ need to be readily available with domestic investments in capacity and capability. Calibrating the clinical and research impact in advance requires an evidence-based framework. The evidence base needed to develop frameworks could be from healthcare outcomes associated with surveillance and modelling, early diagnosis, risk of transmission, vaccine rates and working conditions.

## Methods

We obtained end-user experience through our EPIC mixed methods study to inform the development of a framework [8]. The two-step process allowed the development of a logic model based on real-world evidence by engaging stakeholders within CTU environments and a concept cluster map.

### Engaging stakeholders to construct the framework

Clinical trial unit staff reported cross-sectional and interview data were used to assess the intrusiveness of the system. A key challenge was to identify dimensions that could be used to construct the framework with an index scale. The data gathered was formulated into a logic model as indicated in Figure 1

[INSERT FIGURE 1]

The logic model was designed based on the set measures within the quantitative and qualitative questionnaire of mental health outcomes and opinions of service users, respectively. The yield of results for the logic model also took into account inferences on performance within the UK’s clinical trial networks developing and delivering clinical research associated with communicable and non-communicable diseases.

### Concept map cluster

The concept map was developed based on the evaluation metrics pertaining to the functions and processes that would need to be measured. Contextual knowledge was developed in the form of the EPIC Pandemic Tool which comprises of validated mental health questions to determine depression, anxiety, and burnout. In addition to this, familiarity of the conduct of clinical research within the NHS and academia, knowledge around remote work to setup and deliver studies were included. A thorough grasp on COVID-19 research including ethical and race associated challenges to healthcare professionals and patients were considered [9,10]. The understandings of the operational and logistical challenges to deliver clinical trials were considered in a pandemic setting. The knowledge base for this was used as indicators within the methodological framework. Phase I of the methodological framework transcribed qualitative data and extracted the themes into a data extraction table. The information was analysed, synthesised, and amalgamated with the quantitative data gathered from the EPIC impact Tool using an iterative process.

## Results

Typical logic models are developed from inputs, activities and outputs based on a variety of categories. Our logical model was developed based on spatial features based on the inputs, outputs and outcomes of the EPIC Tool which includes a *cluster* of ideas provides by participants (Figure 2). The understanding of the concepts facilitated the development of the pandemic preparedness framework for CTU staff.

[INSERT FIGURE 2]

### Analysis of qualitative interviews

Six participants consented and completed 1:1 interviews (semi-structured interview script in Appendix One). Interview duration ranged from 26 to 54 minutes (M = 41 minutes). Four women and two men were interviewed, all were white British. Roles represented diversity across the multi-disciplinary team with interview participants including senior statistician, research nurses and trials managers.

All interviews were conducted via secure online facility (password protected teams teleconference), audio-recorded and transcribed in full. Data collection and analysis was integrated with a process based on a framework methodology used to analyse the data including development of a coding frame based on identified key themes and detailed coding of transcripts. Four main overarching key themes were identified, and each was subjected to detailed analyses to identify subthemes and codes with illustrative quotes. The emergent themes are illustrated by verbatims quotes below.

### CTU Job Features

All participants provided some description of their *CTU work* in general terms and independently from the pandemic. Five respondents made statements on the *job stress levels* and identified that “it’s quite a high-pressure job” (P3), “it’s always busy” (P5) but that “clinical trials stress comes and goes but stress tends to fall off when people get a hang of things. Problems come and go. Peaks and troughs” (P1). One participant commented on their *job satisfaction* by pointing out that “I sometimes (enjoy my role). If I have something interesting that is problem solving or stimulating but most of the time, it’s a bit of a grind with lots of documents.” (P2)

Two staff members recognised some *challenges* in their roles - “funding was cut out which is a major issue to bring in new staff. Methodological research isn’t paid for now” (P2); to *change in the public sector* - “in a public sector it would need to set up something new which is quite difficult. People are scared of the concept” (P2); and to the *lack of a shared perspective* - “it also comes back to people managing their own bubbles and not realising what would be useful to other people. So engrossed in the troubles you’ve got, and that’s an age-old problem” (P3). Conversely, a *benefit* that was identified was in relation to the *delocalisation of roles* - “the motivation of geography isn’t an issue anymore and this can be used to feed and fill empty gaps” (P2).

Four participants *suggested improvements* that would, in their opinion, reduce their burden at work. This included *better quality of data* - “[…] better quality of data collection and validation in the cmf. A bit more flexibility in our tool rather than a frozen database” (P2); *better management of timelines* - “[…] if something is going to have a 3 week turnaround that would normally have a 3 day turnaround, just being able to build that in” (P3); a *shorter commute* and interventions around *staff recruitment, retainment and training* - “[…] we had this huge turnover of staff in the past five months and they couldn’t hire anyone, so we had trials and there just wasn’t people, […] the issue we have now is so many people have started but they all need training” (P5).

### Work Life Pre-Covid

All participants offered some contributions on how their work life was before the pandemic. There were differing views about the *office life*, in particular with what concerns *working in a big team/office*. One participant stated that he was part of “a large team and it usually it was fine. People would get on and plug away with their own work. If they needed help or advice then someone was there to help” (P1). However, the same respondent identified that “there were always people that would make everything a drama” (P1). On a similar note, another participant reflected that he “found it quite hard actually to get stuff done. […] it’s too loud and I get distracted really easily. […] I would say there was a lot more inter-personal stress […] I feel like pre-pandemic I didn’t really realise this, but […] I wasn’t perhaps working at the best capacity I could” (P4).

Other participants described their *typical working day* in different terms. One person recognised that they were mostly “in the office, it was quite isolated. It was very small and you only mixed with people from your own trial” (P5), and another respondent highlighted that they “would be in the office every day with a mix of meetings and reading emails. I would sometimes look at data maybe 50% of the time. Not all the meeting were useful.” (P2). Furthermore, one participant identified the impact that a *difficult commute* had on him as “my working day would always shift, I’d work 8-4 so I could try and beat the traffic” (P4).

Finally, when putting *day-to-day activities* into perspective, one respondent reflected that the *possibility to problem solve* was perceived differently pre-pandemic as he felt that although “we knew that we would be expecting curved balls […] nothing felt that there wasn’t a solution.” (P3).

### Work Life during the Pandemic

When asked to describe how their work life changed during the pandemic, participants openly spoke about a range of *challenges* that they had to face and work through. There was consensus that although the workload did not necessarily increase, people at work experienced *increased pressures and lack of staff*. A respondent stated that “finances and grants get over stretched. Indirectly no money to recruit another member of staff” (P2) and that they perceived “more stress as there wasn’t enough staff to help […] out.” (P2) To add to this, some members of staff had to face the issue of *redeployment*, which was described by one participant as “very stressful. Having to go to a new area and work with strangers on something I really didn’t know a lot about” (P6). Additionally, one participant noted that the new circumstances reflected in a *loss of benefits* for CTU staff, such as for instance “pensions being buggered up […] You don’t get to travel to conferences. Getting paid less that someone from a pharma company.” (P2).

At the height of Covid, there was a sense of things being beyond individual control - “it was awful […] your day wasn’t your own […] the adrenalin kicked in because everyone was going into survival mode” (P3). “Mostly things that are out of my control[…] things that are out of my hand that are stressful” (P1). One member of staff highlighted that *finding solutions* became harder as despite having “the same level of enthusiasm towards the things we do, […] some things I have no clue what the solution is now […] nobody knows the answer now.” (P3). This was reflected by participants in managerial roles that acknowledged changes in the way they could *manage people*.

Several respondents noted a number of issues with trial-related work. On one hand, there were simply some activities that could no longer be done. “Some people’s trials were really halted” (P5), “all of the research nurses were redeployed so there was no-one there to open emails or answer queries” (P4). For those trials that could continue, members of staff had to face several *pressures and delays* and come up with *adaptations*. In some instances, it was impossible to obtain necessary approvals, signatures or other paperwork necessary, with one participant commenting that “everyone was experiencing delays across the country […] the industry pretty much shut down. The trial continued, there were things we could do, but there were certain roadblocks that there’s just no getting over.” (P3). Despite this, new Covid-related trials were initiated with associated urgency and other non-Covid trials transitioned to an ‘on-line’ or ‘telephone’ mode of delivery where possible.

Overall, a *level of uncertainty about the future* emerged from participants - “I don’t think things will go back to the way it was.” (P1) and “there is still a degree of uncertainty. What will become the norm and what does a new job look like.” (P2). For one participant, this was also accompanied by a *perceived inability to stop and take a break* - “It has been a journey, we’ve all had our moments. I’m sure someone described it as being on a treadmill and I want to get off but I can’t. I need a break but I can’t get a break.” (P3).

Another cluster of challenges participants had to face were *Covid-related sickness and personal/family consequences* deriving from the pandemic. One person shared their experience of how they “got covid and was really poorly. I was in bed for 3 weeks and it took 4 months to get better. […] I felt incredibly guilty. I was supposed to be doing the trial manager but and I couldn’t.” (P3). Another respondent said “I had covid twice, it was really difficult […] my husband worked in A&E and the nurses were off sick […] he’s had a hell of a time over the last 2 years, he’s lost so many people” (P5).

For most participants this also reflected in the way they had to manage *child- care and family spaces and arrangements*. Many staff members were impacted by “having family around, and having toddlers around” (P3) but also by being “all confined to a very small space” and trying to balance “a five year old to home school and work at the same time” (P6). This in turn meant the traditional ‘working day’ disappearing and being thrust into working extended and often unsociable hours late in the evenings and weekends. The issue of *time boundaries* was perceived differently by participants and there were clearly different expectations from colleagues - “nobody had an escape, nobody had down time but 10, 11 o’clock at night emailing isn’t ideal” (P3) but also “I respond to emails any time e.g., at 11pm I can give a quick answer and it’s done[…] I’m quite responsive with people, I’m not a stickler with hours (P4). “There is an expectation to work more from one person who wants to have meetings at 6-7 in the evening” (P2).

Most of the staff that were interviewed stated that since the start of the pandemic they had to adapt to a new *blended-hybrid and/or remote way of working* and experienced a change in their working patterns and base. Five participants mentioned mixed experiences in relation to having to *reorganise their spaces at home* to facilitated their work - “I first was using the dining room as I hoped it wasn’t going to be that long. […] I moved upstairs into the spare room. I have a full set up now … a mini version of my office” (P1). “We were in the middle of a house extension and we were living in our kitchen. […] I was sitting on a little chair on the floor in my kitchen at the time or sitting on my bed upstairs to work. So that didn’t help really.” (P6). This also translated into a slight dispersal of the workforce as some participants were continuing to work from home, some in the office and others working a hybrid between home and the office - “there aren’t many people showing up to work in the office it feels pointless to go in” (P1).

This transition also came with its own *IT and technical issues* - “this hybrid model, especially IT functions, is incredibly difficult. You’d go in and somebody has taken your mouse and your keyboard for example.” (P3) and “I had to use my home computer which was very difficult. I had access issues for confidential documents” (P5). Participants had mixed views about moving from face-to-face meetings to *online meeting* platforms during the pandemic. “I used to have two meetings a week on zoom and there was a time when there were 7 a day. That’s no good for anyone. No time to go for a wee.” (P3). “You bounce from meeting to meeting rather than doing it physically” (P1). There were positives as people were “able to jump into a meeting easily.” (P2) and another person reflected that “I do think your true self comes out, everyone’s much more relaxed in a zoom environment than we were before […] its taken the pressure off.” (P3).

Additionally, the pandemic was a time where continuity and communication became very difficult - “The biggest problem was in the communication […] We realised no-one’s coming back full-time and had to adapt to different ways of working and communicating” (P5). In response to this, some teams made efforts to ensure a team spirit continued throughout by organising online social events to *keep people connected*. The majority of participants did however find benefits in the level of *flexibility and choice* that the new arrangements brought along - “I get better results out of people if I’m more flexible with hours. It’s give and take” (P4). Two respondents seemed to particularly benefit from the new arrangements in terms of their *work-life balance* as they both had systems in place that would grant them the *ability to disconnect* between home and work. This way of working made it easier for them to *take care of both home and work commitments*. One participant shared that the downside to this was the feeling of isolation - “It was a bit lonely at home, not seeing anyone…quite lonely.” (P6)

There was consensus among respondents that the new ways of working resulted in *increased efficiency, quality, and productivity*. One member of staff noted that “things have become more efficient really […] No distractions.” and that “now you have a bit more spare time and are not being pestered by colleagues all of the time in the office it helps.” (P2). This was echoed by other participants who felt that “people [are] being more responsible for their own workloads. Getting more out of people.” (P3) and “I feel like I have a higher stress level but I’m more focused and productive. I am able to work more efficiently and better from home. They get so much more from me.” (P4).

Some participants commented on the perceived *higher levels of flexibility, acceptance and tolerance towards and from others*. One respondent noted that even though it was not “all plain sailing […] we understand each other better. We understand humans and what humans need better within our team […] there’s a level of acceptance now.” (P3). Another participant said that “it’s a lot more understanding and flexible but we’re all quite chilled out here I think” (P6). In connection with this, one staff member found that there is now a higher appreciation of the *key role people play in CTU* which, in turn, led to a *higher commitment from the team, increased cohesion and better relationships in the team* and *increased support for people*. This was in accordance with other two respondents, who felt that “there’s always support as a unit they have been really good across the unit […] we actually collaborated really well as a unit” (P5) and “I feel really well supported by my team. We’re working in a way that works for you and your manager.” (P4).

*New Normal – Living with Covid*

Participants described their new *work-related stress* levels between a 3 and a 6. Two participants felt things were worse since Covid, whilst two said it was better, one that things had returned to *pre-Covid levels*. Views diverged on the contributors to this - “given the changing working conditions” (P2), “working from home has taken away loads of stress” (P4), “my study has restarted, and we’re also still involved in covid studies as well” (P6), “my study now is significantly more complicated than the one I was running through covid” (P4). Overall, there was a shared understanding of how different individuals might have had *different experiences*, with regards to adjusting to the living situation in the midst of the pandemic and as part of the ‘new normal’.

Participants shared a sense of uncertainty and anxiety surrounding *returning to the office* and there were mixed views on the desirability of *getting back into a routine*. “Technically we’re back but not everyone is yet. All the desks have moved. Having to travel again is weird. […] I think the anxiousness is around getting trains again, walking again, meeting people again, all in like just being in a social situation. […] I’m a very routine person so now I’m back in a routine I’m actually loving it […]Some people left because they don’t want to be back in the office. […] I didn’t realise till I started to go back in just how refreshing it is to have, like, even a laugh with someone in your office.” (P5). “We have been [more anxious] in on occasion. People don’t enjoy the thought of it [going into the office] but the human element and I know some people absolutely need it.” (P3).

However, the majority of participants felt safe in the office knowing there were new safety regulations in place - “there is lots of PPE dispensers and reduced capacity and we’re only in 2-3 times a week […] not the full staff anyway.” (P1). “I don’t feel anxious within the premises of work. I feel safe and have had the vaccines so the direct threat to myself isn’t an issue” (P2).

Finally, it emerged from interviews that a lot of the changes and *new processes* that were implemented during the pandemic would remain in place as part of the ‘new reality’ - “There’s a lot more applications we have to fill in. Research applications, basically you’ve got to apply to work, to use any anywhere and get approval, use the clinic rooms for studies. That’s a little bit, but that’s just new normality” (P6).

### Strategic Framework for CTU Workforce

The below strategic framework has been derived from the above thematic analysis to explore five core pillars for the CTU workforce. As explored within the *Work Life during the Pandemic* theme, the Operational Pillar, Innovator Pillar, and Ecosystem Pillar are crucial to the activities and efficacy of the CTU workforce. Ensuring better training and enhancing effective communication for emergency scenarios, such as a pandemic, would enable for better wellbeing and operations within the CTU team. This will enable for better managements of teams and workflows in abnormal working situations. The Governance Pillar is core to the CTU work, regardless of the working situations and this should be maintained. The Economic Pillar is more relevant to the higher-order positions within the CTU workforce.

## Discussion

Based on the qualitative findings, it appears, there is significant variability in experiences and opinions around *lived experience* professionally and personally during the pandemic. Participants reflected on and provided powerful insights into the impact of Covid-19 whilst working to deliver research in a CTU environment throughout the pandemic. Staff were exposed to a wide range of new and distressing circumstances, which highlighted the importance for managerial and supervisory staff to ensure that workers are appropriately supported at all times. This becomes ever more important in exceptional and unprecedented working and life circumstances, such as those that manifested during the Covid-19 pandemic. Moreover, Covid-19 has demonstrated the importance of diversity and the disproportionate impact that minority populations endured [10, 11]. Research demonstrated that the pandemic has had a unique and negative impact on minority ethnic staff within the NHS [12]. Around 45% of NHS workers are from ethnic minority groups however between 60-75% of healthcare worker deaths due to the pandemic were of these minority staff members [10,12]. This suggests that ethnicity is a significant contributory factor to the impact of Covid-19 on ethnic minority staff, but this area remains under-researched and not well-understood [12]. Based on our study results, there appears to be under representation of ethnic minorities in CTU staff. Adequate representation could have facilitated more meaningful conclusions about the impact of Covid-19 on minority ethnic CTU staff within this study.

It has been recommended that all staff should be closely monitored post-pandemic, especially those groups at higher risk (e.g., minority ethnic groups or older staff), to ensure early recognition of and adequate support for any arising problems [13]. Supporting healthcare workers and promoting their psychological wellbeing is vital at minimising the risk of mental health conditions and will enable them to overcome the struggles faced throughout the pandemic [13]. A variety of efforts have been suggested in order to prepare for future pandemics. Increased availability of pharmaceutical interventions, such as antivirals or vaccines, could help reduce duration of illness and reduce the spread of infection [14,15]. However, it would not be enough to solely increase availability of medicines if pharmaceutical commodities are widely unavailable, which seems to be the case in many developing countries [15]. Needles, syringes, and temperature-controlled storage units are some of the basic necessities which should be delivered to healthcare facilities across the globe [14,15].

Infection and mortality rates drastically increase throughout the pandemic, with healthcare services falling under immense pressures [15]. All countries remain unequipped to deal with such a high patient influx in regard to staff, hospital beds and pharmaceutical equipment [16]. In order to ensure a global standard for pandemic preparedness it is crucial for global governing bodies, such as the World Health Organisation (WHO), to develop comprehensive guidance and protocols [17]. Whilst the WHO have released a national preparedness checklist, it only describes generic approaches, not accommodating for developing countries which may lack facilities [15,17]. Models that forecast impacts of pandemics and accordingly suggest feasible interventions and guidelines would be more useful [15]

Such findings can be used to formulate various guidelines and frameworks which can be implemented in the CTU workforce. It can be used to facilitate and develop economic models which can be used at an organisational and also national level. In 2019, clinical research generated £2.7 billion to the UK economy, creating almost 50,000 full-time job positions [18]. For the NHS more specifically, clinical trials generated £355 million, saving the NHS £28.6 million in costs [18]. Further research into CTU frameworks can help to understand which operations mechanisms work best in order to sustain and bolster these economic outputs. This would feed into the operational and governance structures within CTU work. Ensuring alignment and efficiency in procedures through standardised would further escalate positive outcomes.

## Limitations

Despite considerable challenges in recruitment to this interview study, the depth of data from the included participants reflected very different experiences across CTU staff and provided some powerful insights into the impact of COVID on professional lives in the context of very personal experiences. Due to the small sample size, it is difficult to generalise findings to the broader CTU network. Experiences reported by participants reflected the diversity of human nature and coping. Enrolment of participants could have been increased with less pressure on the CTU workforce. Furthermore, there were numerous observational, and vaccine related interventional studies during this period that could have resulted in this study competing with other portfolio studies for staff recruitment and participation. Cultural adaptations could have been considered when conducting further work based on the evidence gathered within this study if ethnic minorities were better represented. There were considerable challenges to increase participants for the qualitative component of the study. Longitudinal data collection could be a useful step to continue to assess these findings to aid CTU workforce and employers make quality improvements.

## Conclusion

Our study indicates the substantial personal impact the CTU workforce have encountered during the pandemic. Overall, participants felt well-supported where needed and had developed a resilience to what had been some very challenging situations. Most people however experienced a variety of challenges with the sudden changes linked to home working. The viability of sustainable clinical trial conduct is based on multi-professional involvement in CTU settings thus, it would be in the interest of all healthcare professionals to improve the support systems available to better manage working conditions especially as part of pandemic preparedness.

Recommendations to inform ‘Future Preparedness’ in supporting the CTU workforce in delivering pandemic and non-pandemic research include ensuring continuity and clarity of communication via different media, providing opportunities for flexibility in working hours, within reasonable constraints, to ensure staff are not pressured into work during times they would not usually do so. Additionally, it would be appropriate to provide opportunities for collaborative problem-solving via different media as per the needs of individual team members and to maintain consistency, where possible, in frequency and content of contact so that staff feel supported in different aspects of their working roles. Finally, it would be appropriate to complete a contingency planning of supplies of necessary equipment for effective home working, which should be considered in line with all relevant health and safety legislation to ensure the wellbeing of CTU workforce as they lead in the delivery of cutting edge innovative clinical trials that will be tomorrow’s interventions.

## Data Availability

All data produced in the present work are contained in the manuscript

## Acknowledgements

The authors acknowledge support from Southern Health NHS Foundation Trust.

